# Prevalence, determinants, and trends in the experience and perpetration of intimate partner violence among a cohort of gay, bisexual, and other men who have sex with men in Montréal, Toronto, and Vancouver, Canada (2017-2022)

**DOI:** 10.1101/2023.07.24.23293112

**Authors:** Stephen Juwono, Jorge Luis Flores Anato, Allison L. Kirschbaum, Nicholas Metheny, Milada Dvorakova, Shayna Skakoon-Sparling, David M. Moore, Daniel Grace, Trevor A. Hart, Gilles Lambert, Nathan J. Lachowsky, Jody Jollimore, Joseph Cox, Mathieu Maheu-Giroux

## Abstract

**Purpose:** Longitudinal data on the experience and perpetration of intimate partner violence (IPV) among gay, bisexual, and other men who have sex with men (GBM) is limited. We estimated the prevalence of past six-month (P6M) physical and/or sexual IPV (hereafter IPV) experience and perpetration, identified their determinants, and assessed temporal trends, including the impact of the COVID-19 pandemic.

**Methods:** We used data from the Engage Cohort Study (2017-2022) of GBM recruited using respondent-driven sampling in Montréal, Toronto, and Vancouver. Adjusted prevalence ratios (aPR) for determinants and self-reported P6M IPV were estimated using generalized estimating equations, accounting for attrition (inverse probability of censoring weights) and relevant covariates. Longitudinal trends of IPV were also assessed.

**Results:** Between 2017-2022, 1,455 partnered GBM (median age 32 years, 82% gay, and 71% white) had at least one follow-up visit. Baseline proportions were 31% for lifetime IPV experience and 17% for lifetime perpetration. During follow-up, P6M IPV experience was more common (6%, 95%CI: 5-7%) than perpetration (4%, 95%CI: 3-5%). Factors associated with P6M IPV experience include prior IPV experience (aPR=2.79, 95%CI: 1.83-4.27), less education (aPR=2.08, 95%CI: 1.14-3.79), and substance use (injection aPR=5.68, 95%CI: 2.92-11.54, non-injection aPR=1.70, 95%CI: 1.05-2.76). Similar factors were associated with IPV perpetration. IPV was stable over time; periods of COVID-19 restrictions were not associated with IPV changes in this cohort.

**Conclusion:** Prevalence of IPV was high among GBM. Determinants related to marginalization are associated with an increased risk of IPV. Interventions should address these determinants to reduce IPV and improve health.

## Introduction

Intimate partner violence (IPV) encompasses harmful behaviors by a former or current intimate partner and includes psychological, physical, and sexual violence.^1^ IPV can have long-lasting physical, mental health, and economic consequences.^2,3^ Gay, bisexual, and other men who have sex with men (GBM)^4,5^ may experience IPV in their relationships; limited data from 11 countries, including Canada,^4^ indicate that up to 33% of GBM reported any lifetime IPV experience and 29% reported any IPV perpetration.^6^

Despite recent efforts to better describe and understand IPV among GBM, there is a paucity of longitudinal research on the topic.^6–9^ The few existing studies have focused on the specific transition period spanning adolescence to young adulthood^10–13^ and only one reported longitudinal IPV prevalence estimates, with a focus on dating violence.^13^ Overall, this work suggests that the prevalence of IPV experience and perpetration among GBM may be relatively stable over time.^10–13^

This scarcity of longitudinal research among GBM hampers our understanding of risk factors and consequences associated with IPV in this group. Although previous cross-sectional research has highlighted risk factors for IPV within a broader syndemic context,^14–16^ wherein negative health states synergistically contribute to each other and exacerbate IPV, it remains uncertain whether these associations persist when evaluating IPV over time. Among cross-sectional studies, risk factors for IPV among GBM include socio-demographic characteristics (e.g., ethnicity, age),^15,17,18^ health-related factors (e.g. anxiety, depression, substance use),^19,20^ and contextual factors (e.g. sexual minority status, partnership dynamics).^21,22^ Addressing IPV among GBM requires not only learning from successful women-centered services and interventions,^23,24^ but also developing novel services and interventions designed specifically for these communities.^25^

Temporal trends in recent IPV prevalence might have been affected by COVID-19 disruptions, such as physical distancing and lockdown measures.^26^ Further, IPV-prevention and - mitigation resources were scaled-down during the COVID-19 pandemic.^27,28^ The impact of COVID restrictions on IPV among GBM is unclear as research is limited,^29^ however studies among women suggests that both the incidence and severity of IPV may have increased during the pandemic.^30,31^ Thus, we expect similar increases in IPV among GBM during periods of COVID-19 restrictions. Studying the impact of public health pandemic measures on IPV experience and perpetration would help enhance our understanding of the burden and nature of IPV among GBM.

Using data from a large, population-based, longitudinal cohort study of GBM in Canada’s three largest cities (Montréal, Toronto, and Vancouver), this investigation sought to estimate the prevalence of recent physical and/or sexual experience and perpetration of IPV, investigate longitudinal determinants of IPV, and assess temporal trends in IPV experience and perpetration in relation to the COVID-19 pandemic.

## Methods

### Data and study design

The *Engage Cohort Study* (Engage) is a longitudinal study of GBM (2017-2022) recruited between February 2017 and August 2019 via respondent-driven sampling (RDS) in Montréal, Toronto, and Vancouver.^32,33^ RDS is a modified chain-referral method that can be used to obtain a more representative sample of hidden populations.^34^ Initial participants, or “seeds”, were purposively selected to characterize diverse features of the GBM population in Canada. Eligibility criteria for enrolment included being aged ≥16 years old, identifying as a cisgender or transgender man, reporting sex with another man in the past six months (P6M), and having the ability to read English or French. Follow-up visits were scheduled at 6-month intervals across the three cities, except for the first two years in Montréal and Toronto, where study visits occurred every 12 months. After providing written informed consent, participants self-completed questionnaires on socio-demographic characteristics, health and healthcare use, community and societal context, partnership characteristics, experience and perpetration of IPV, as well as sexual behaviors and substance use. A $50 honorarium was provided after each completed visit and $15 was provided for each participant they recruited into the study (up to 6).

### Analytical sample

We included participant data collected between February 2017 and August 2022 and only considered visits if the participant reported being in a relationship with a primary partner at the index visit or the previous one. Participants without follow-up visits were excluded from our sample. Partnership status was assessed based on whether the participant stated having a “relationship with a primary partner” or reported their primary relationship status as either dating, common-law, or married. This was motivated by our focus on IPV, as opposed to dating violence. Figure S1 presents the exclusion flow diagram.

### Measures

#### Intimate partner violence

Our primary outcomes of interest were the prevalence of P6M physical and/or sexual IPV (hereafter referred to as IPV) experience and perpetration. Given the lack of consensus on appropriate measures to operationalize a definition for psychological IPV,^1,9^ we focused on physical and sexual IPV. For all IPV questions, a modified version of the Revised Conflict Tactics Scale^16,22^ was used to obtain information on the baseline lifetime experience and perpetration of physical and sexual IPV, and, for the follow-up visit, experience and perpetration in the P6M (exact wording in Appendix I(a)). Baseline IPV was included to evaluate *de novo* or new IPV at follow-up visits. Questions on P6M IPV were not included in the first follow-up questionnaire in Montréal and Toronto.

Covariates from the following domains were selected to examine determinants of IPV: socio-demographic, HIV status, partnership characteristics, childhood sexual abuse, sex work, anxiety and depression, problematic alcohol use, and unregulated substance use. These variables have been previously identified as being associated with IPV experience or perpetration among GBM partnerships.^16,17,35,36^ Refer to Supplementary Table S1 for detailed definitions on the selected covariates.

### Statistical analyses

All proportions and analyses, unless otherwise specified, were adjusted using RDS-II weights,^37,38^ which are inversely proportional to the participants’ reported network size. Descriptive statistics, including lifetime IPV experience and perpetration at baseline, were calculated to examine characteristics, stratified by city.

As part of the descriptive analysis, we illustrated the overall city-specific temporal trend of P6M IPV experience and perpetration via a natural cubic spline regression model with robust standard errors (i.e., generalized estimating equations, GEE). Final knot locations were chosen using the quasi-likelihood model criterion (QIC).^39^ A bootstrap method was employed to construct confidence intervals (CIs). We resampled study participants with replacement to address within-participant correlation, repeating this process 1,000 times.

Data from each city were pooled to calculate crude and inverse probability of censoring-weighted prevalence ratios (PRs) for the association between P6M IPV and covariates. To account for autocorrelation between visits, we used Poisson regression models with robust standard errors (GEE). Adjusted prevalence ratios (aPR) were estimated by controlling for age, city, sexual orientation, gender, ethnocultural group, and educational level for both experience and perpetration as the covariates were largely time-invariant. Inverse probability of censoring weights (IPCW) was used in all analyses to account for lost to follow-up. RDS-II weights were not included in regression models as they are often unnecessary to obtain unbiased estimates.^38,40^

Missing observations for IPV outcomes were dropped from bivariate and multivariable analyses under the assumption that missingness is not informative of IPV. Individuals with missing data for covariates were included in all analyses using the missingness indicator method.^41^

Because of the wide variation in how psychological IPV is conceptualized, we conducted a sensitivity analysis which included questions on verbal violence to assess whether a broader definition of IPV would impact our results. All analyses were conducted in R (Version 4.1.0, R Foundation for Statistical Computing, Vienna, Austria) using the packages *RDS, geepack, survey, splines.*^42–45^ A full list of packages used can be found in Appendix I(c). Results are presented according to STROBE-RDS guidelines^52^ (Supplementary Table S2).

### Ethics approval

The Engage study was approved by the research ethics boards of each principal investigators’ respective institutions.

## Results

### Sample characteristics

Our analysis included 1,455 partnered GBM with at least one follow-up visit: 740 (51%) in Montréal, 280 (19%) in Toronto, and 435 (30%) in Vancouver. Altogether, participants contributed 6,537 observations between February 2017 and August 2022 (Table 1). The median number of follow-up visits was 4 (Interquartile range, IQR: 3-4). At baseline, the median age across cities varied between 30 and 32 years, with most participants identifying as cisgender men (94%), gay (82%), English-, French-Canadian, or European (71%); Canadian-born (66%); and having a post-secondary education (87%). Throughout the study period, the overall proportion of GBM in a relationship at any given visit ranged between 68% and 98%, and between 66% and 86% reported having a primary partner, with 30% to 65% cohabiting with their partner at any given time.

**Table 1.**
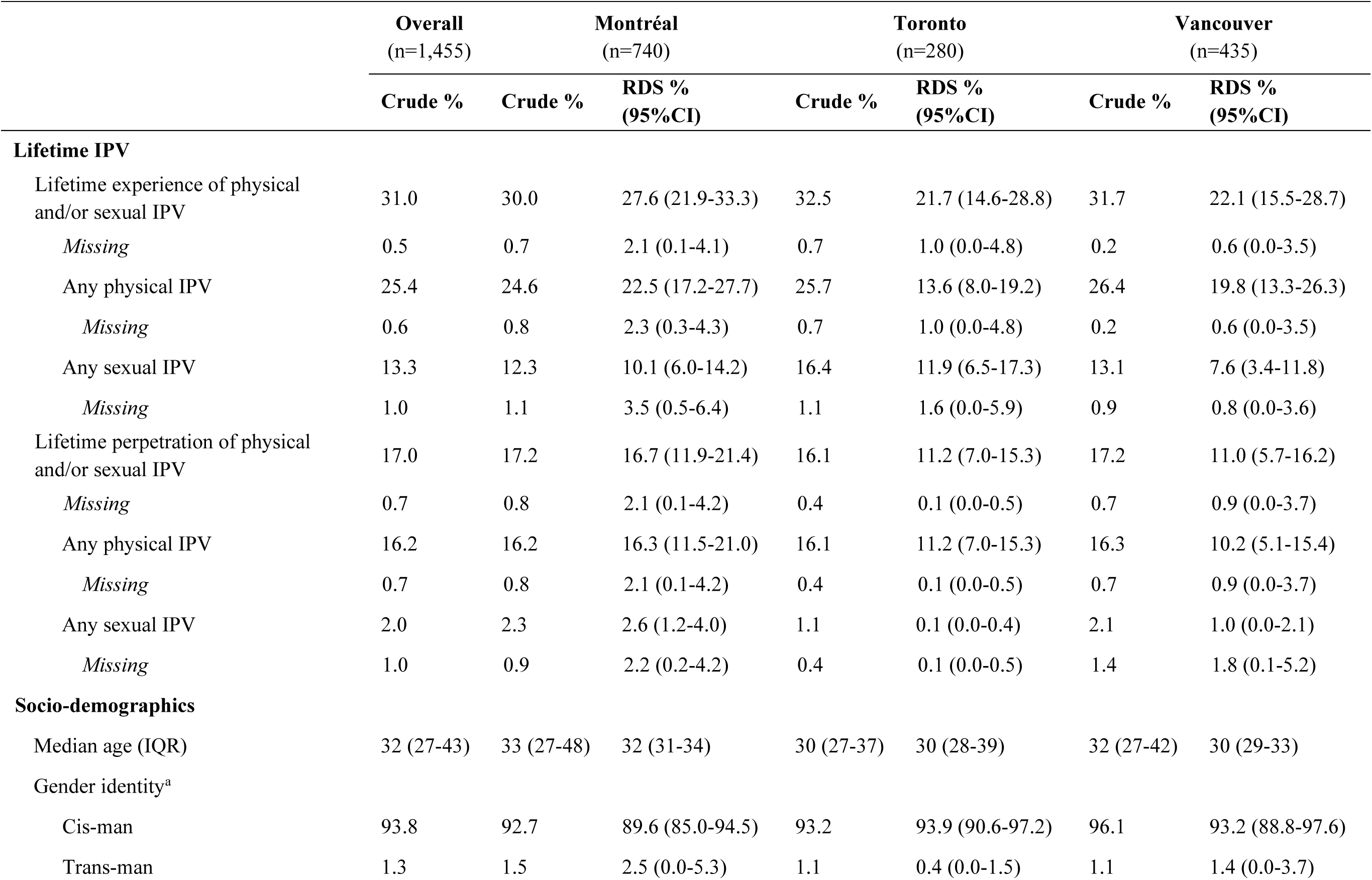
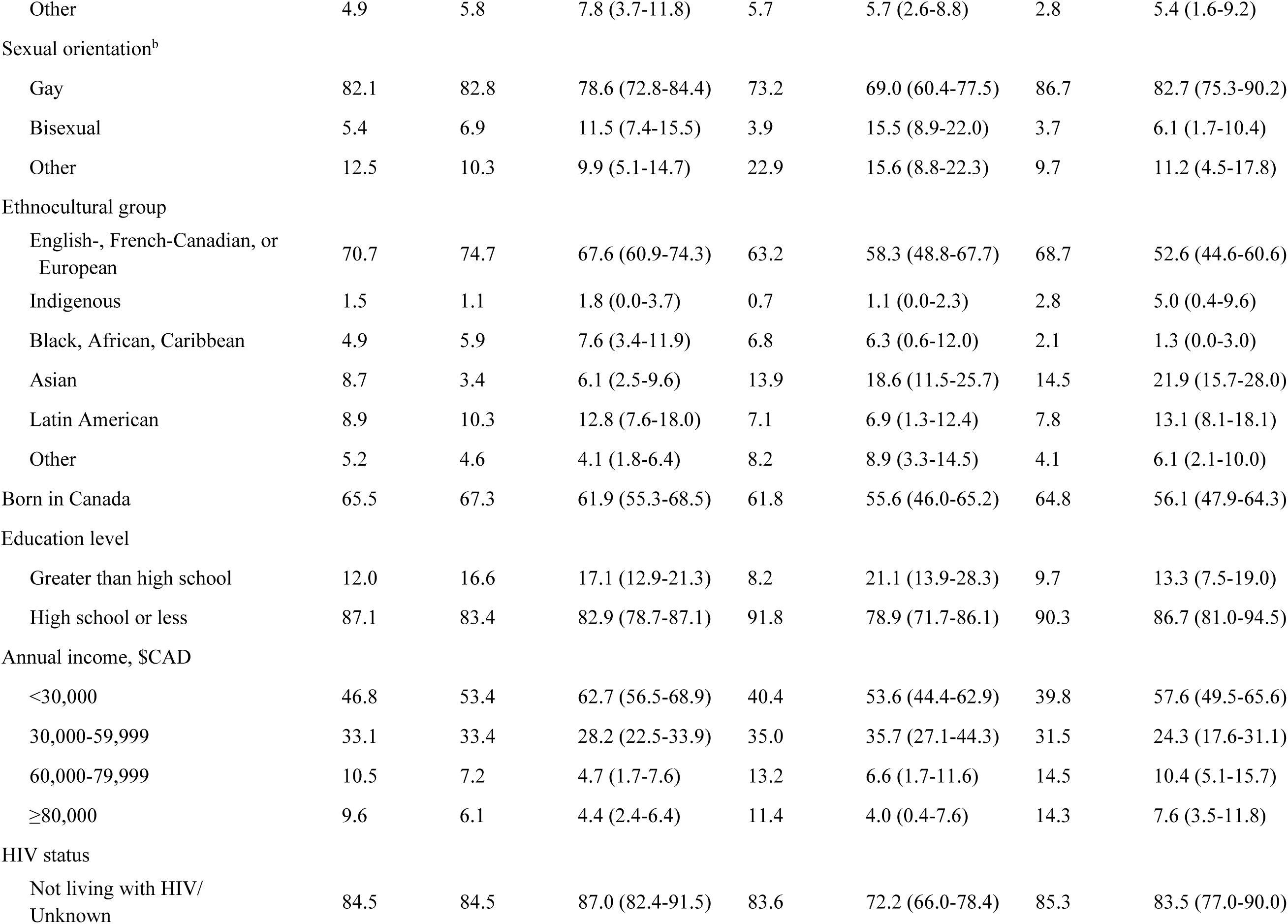
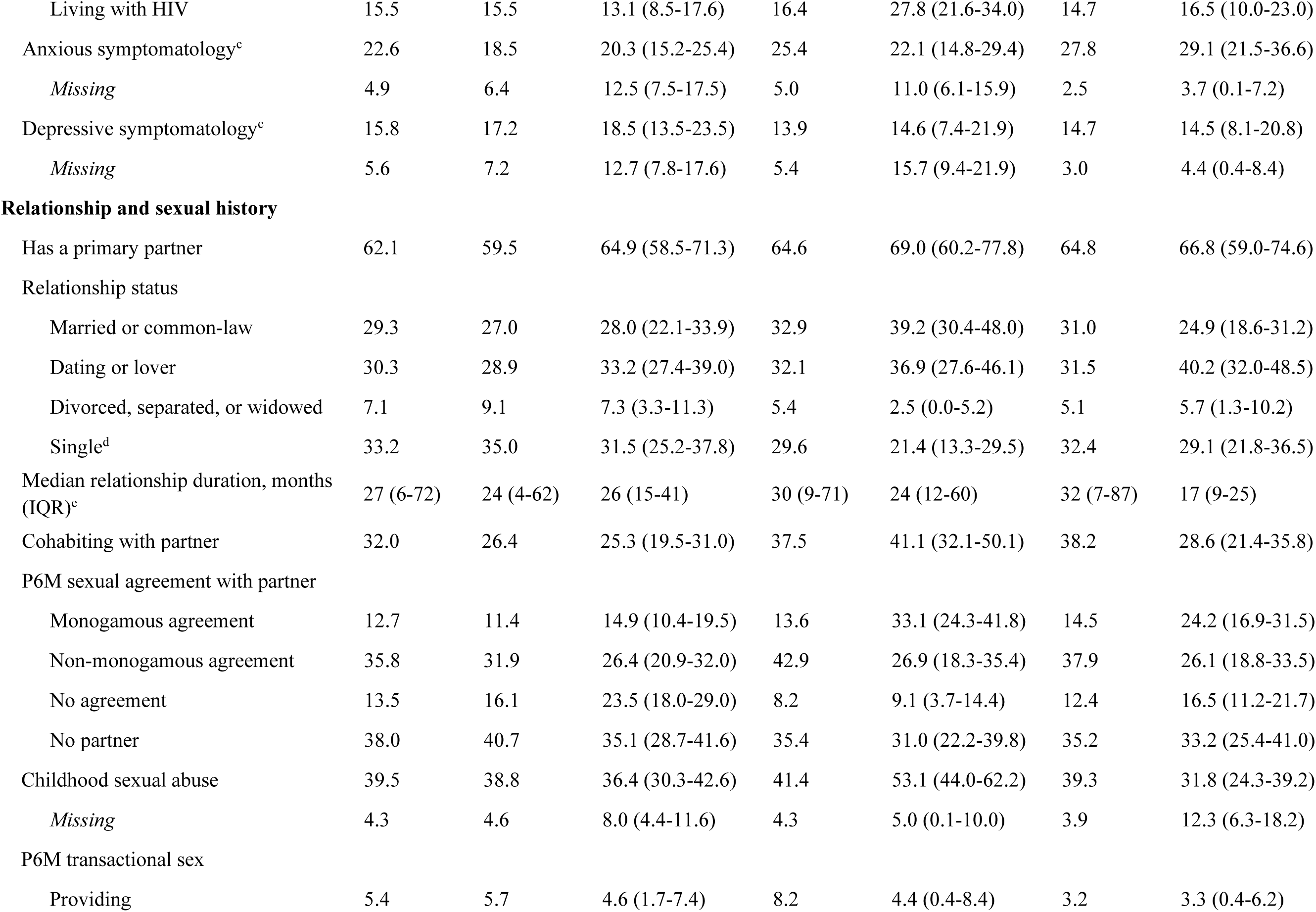
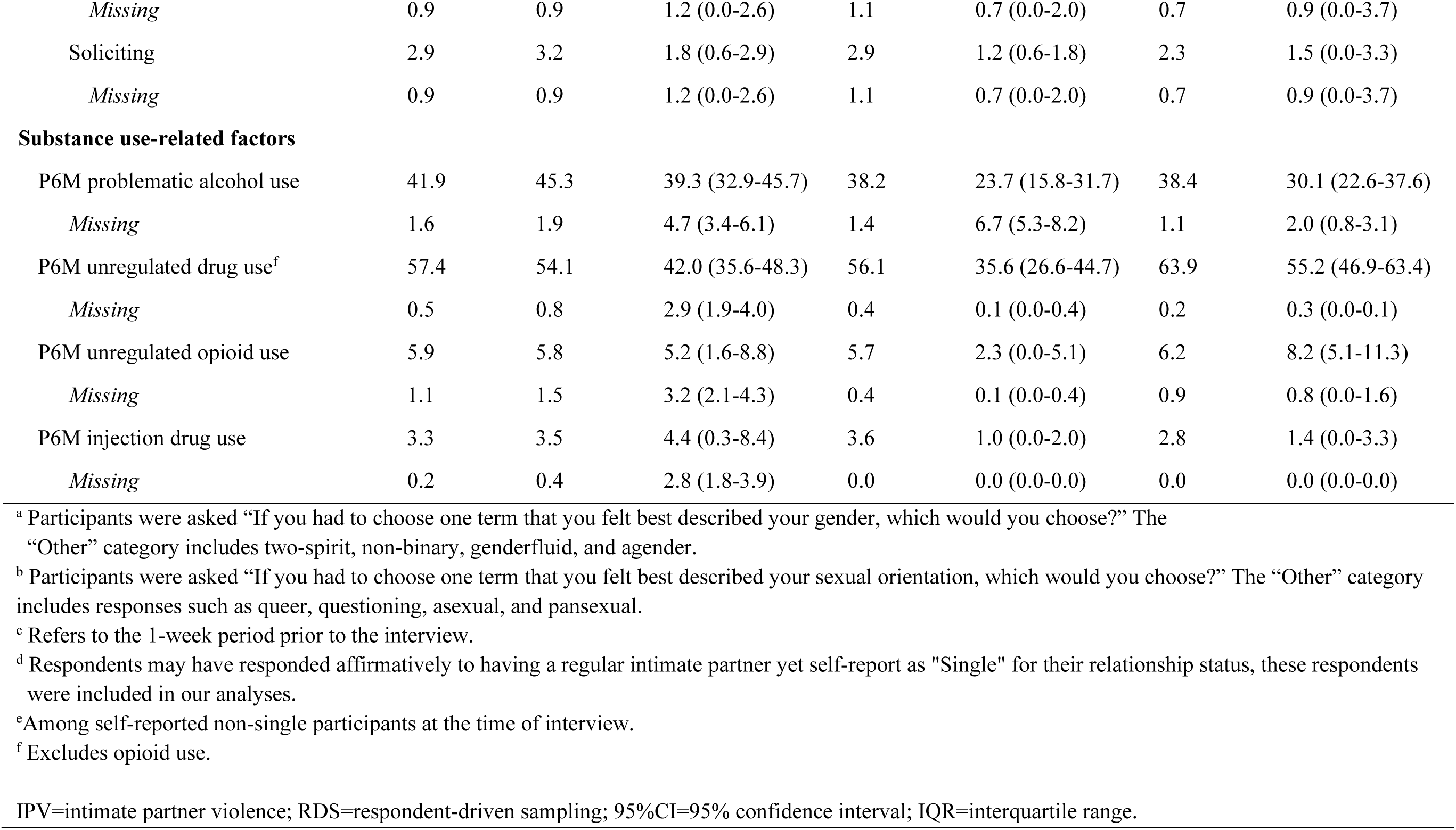
Experience and perpetration of intimate partner violence and baseline characteristics of partnered *Engage* participants with follow-up in Montréal, Toronto, and Vancouver (crude and respondent-driven sampling-adjusted, 2017-2019).

At baseline, the proportion of participants reporting prior IPV experience was 31% (95% CI: 29-33%), while those reporting perpetration was lower (17%, 95% CI: 15-19%), with similar results in each city for crude and RDS-adjusted estimates (Table 1). Lifetime experience of physical IPV (25%; 95% CI: 23-28%) was more commonly reported than that of sexual IPV (13%; 95% CI: 12-15%). Lifetime reporting of perpetration of physical IPV was 16% (95% CI: 14-18%), and that of sexual IPV was 2% (95% CI: 1-3%). Lifetime reporting of IPV experience and perpetration were comparable across cities. The proportion of participants that reported both lifetime experience and perpetration of IPV was 13% (95%CI: 11-15%).

### Incident experience and perpetration of intimate partner violence

During any follow-up visit, 7% (*n* = 85) of participants reported P6M IPV experience, and 4% (*n* = 51) reported P6M IPV perpetration (Table 2). Differences in follow-up duration between those with exposure to P6M IPV experience or perpetration and those without, as well as differences between cities, were minimal. During follow-up, 13% of participants had missing information on IPV. Of the 7% and 4% reporting P6M IPV experience or perpetration, almost 50% was new and among participants who had never previously reported IPV (Table 2).

**Table 2.**
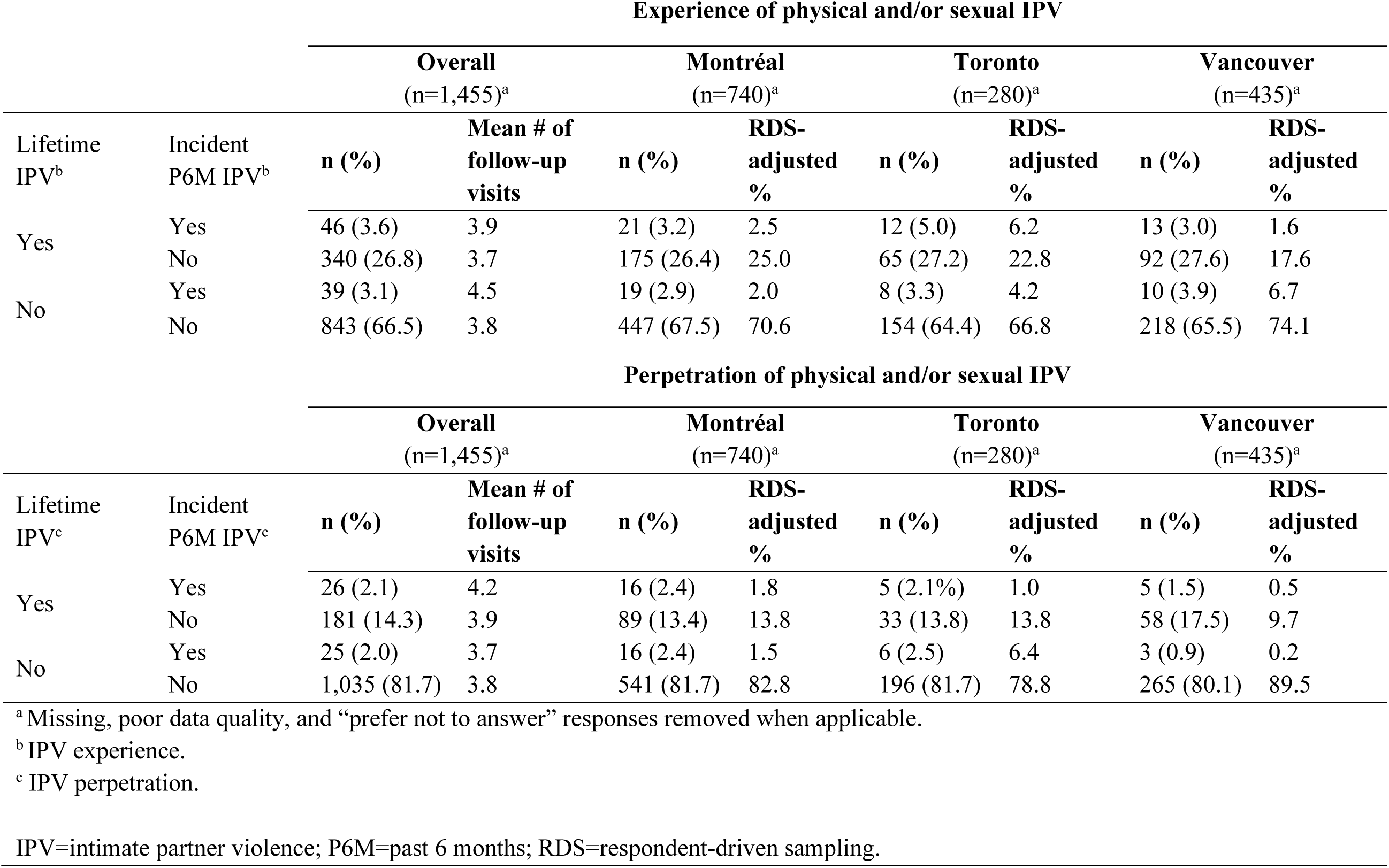
Incidence of self-reported physical and/or sexual intimate partner violence among partnered *Engage* participants with follow-up in Montréal, Toronto, and Vancouver (crude and respondent-driven sampling-adjusted, 2017–2022).

### Determinants of intimate partner violence experience in the past 6 months

Reports of experience of IPV at baseline were associated with P6M experience of IPV during follow-up (adjusted prevalence ratio [aPR] 2.79; 95% CI: 1.83-4.27). The same was true for perpetration (aPR 2.46, 95% CI: 1.51-4.01). Overall, there was a positive association between the risk of reporting P6M IPV experience and being part of a vulnerable group, including those of younger age, lower education level, and those who use unregulated (injection and non-injection) substances (Table 3). In addition, participants who identified as Indigenous, were HIV seropositive, belonged to the lower income group, those with anxious symptomatology, history of childhood sexual abuse (CSA), and those having no sexual agreement with their partner, were associated with increased risk of P6M IPV experience (Table 3).

**Table 3.** Associations between socio-demographic, partnership characteristics, mental health, and substance use, and experience or perpetration of past 6 months physical and/or sexual intimate partner violence (IPV) during follow-up among partnered *Engage* participants in Montréal, Toronto, and Vancouver (crude and adjusted, 2017–2022).

|  | Physical and/or sexual IPV experience<br>in the past 6 months<sup>a</sup> |  | Physical and/or sexual IPV<br>perpetration in the past 6 months<sup>a</sup> |  |
| --- | --- | --- | --- | --- |
|  | PR (95% CI)<sup>b</sup> | aPR (95% CI)<sup>b, c</sup> | PR (95% CI)<sup>b</sup> | aPR (95% CI)<sup>b, c</sup> |
| Reported physical and/or sexual IPV experience at baseline | 2.90 (1.83-4.60) | 2.79 (1.83-4.27) | 3.04 (1.66-5.57) | 2.78 (1.54-5.02) |
| Reported physical and/or sexual IPV perpetration at baseline | 2.79 (1.73-4.50) | 2.46 (1.51-4.01) | 6.60 (3.70-11.79) | 6.11 (3.46-10.79) |
| Age |  |  |  |  |
| < 30 | 1.59 (0.77-3.29) | 1.62 (0.78-3.36) | 1.18 (0.49-2.86) | 1.44 (0.56-3.71) |
| 30-44 | 1.86 (1.03-3.34) | 2.02 (1.13-3.60) | 1.49 (0.73-3.05) | 1.86 (0.88-3.95) |
| 45+ | Referent | Referent | Referent | Referent |
| City |  |  |  |  |
| Montréal | Referent | Referent | Referent | Referent |
| Toronto | 1.58 (0.89-2.82) | 1.82 (1.00-3.32) | 1.01 (0.49-2.06) | 1.51 (0.75-3.04) |
| Vancouver | 0.95 (0.55-1.62) | 1.20 (0.69-2.09) | 0.49 (0.21-1.14) | 0.67 (0.29-1.59) |
| Sexual orientation |  |  |  |  |
| Gay | Referent | Referent | Referent | Referent |
| Bisexual | 2.21 (1.01-4.86) | 1.77 (0.85-3.69) | 3.11 (1.49-6.48) | 2.73 (1.25-5.98) |
| Other | 1.50 (0.82-2.73) | 1.20 (0.64-2.28) | 1.26 (0.57-2.81) | 1.17 (0.46-3.00) |
| Gender |  |  |  |  |
| Cisgender man | Referent | Referent | Referent | Referent |
| Other<sup>d</sup> | 1.35 (0.65-2.82) | 0.85 (0.37-1.97) | 1.11 (0.38-3.18) | 0.85 (0.37-1.97) |
| Ethnocultural group |  |  |  |  |
| English- or French-Canadian/European | Referent | Referent | Referent | Referent |
| Aboriginal or Indigenous | 3.02 (1.23-7.40) | 2.99 (1.25-7.15) | 3.02 (0.85-10.68) | 2.99 (1.25-7.15) |
| Asian | 0.71 (0.26-1.93) | 0.59 (0.24-1.44) | 0.13 (0.02-0.97) | 0.59 (0.24-1.44) |
| Latin American | 0.81 (0.35-1.87) | 0.81 (0.36-1.84) | 0.37 (0.11-1.22) | 0.81 (0.36-1.84) |
| African/Caribbean/Black | 0.66 (0.21-2.11) | 0.60 (0.19-1.85) | 1.44 (0.52-3.99) | 0.60 (0.19-1.85) |
| Other | 1.04 (0.47-2.29) | 1.10 (0.51-2.38) | 0.49 (0.12-2.09) | 1.10 (0.51-2.38) |
| Born outside Canada | 0.81 (0.48-1.35) | 0.97 (0.53-1.79) | 0.65 (0.33-1.25) | 1.03 (0.46-2.30) |
| High school education or less | 2.10 (1.20-3.70) | 2.08 (1.14-3.79) | 3.33 (1.71-6.49) | 2.08 (1.14-3.79) |
| Annual income, \$CAD | | | | |
| <30,000 | 2.37 (1.25-4.49) | 2.27 (1.22-4.24) | 2.20 (0.83-5.84) | 1.72 (0.65-4.53) |
| 30,000-59,999 | 1.75 (0.91-3.35) | 1.72 (0.90-3.27) | 2.14 (0.84-5.41) | 1.88 (0.73-4.87) |
| 60,000-79,999 | 1.76 (0.87-3.57) | 1.71 (0.84-3.47) | 1.79 (0.62-5.13) | 1.69 (0.59-3.61) |
| ≥80,000 | Referent | Referent | Referent | Referent |
| Living with HIV | 1.61 (0.93-3.57) | 1.94 (1.04-3.59) | 1.44 (0.67-3.09) | 1.46 (0.59-3.61) |
| Relationship Status |  |  |  |  |
| Married or common-law | 0.61 (0.34-1.10) | 0.64 (0.36-1.15) | 0.80 (0.37-1.72) | 0.92 (0.43-1.95) |
| Dating or lover | 0.75 (0.47-1.19) | 0.74 (0.47-1.18) | 0.99 (0.52-1.87) | 1.00 (0.54-1.86) |
| Single, divorced, separated, or widowed | Referent | Referent | Referent | Referent |
| Cohabiting with partner | 1.07 (0.67-2.73) | 1.10 (0.69-1.77) | 1.04 (0.61-1.77) | 1.16 (0.68-1.98) |
| Relationship duration, months |  |  |  |  |
| ≤ 6 | 1.30 (0.74-2.26) | 1.18 (0.67-2.07) | 1.21 (0.61-2.42) | 0.99 (0.50-1.98) |
| 7-12 | 1.67 (0.83-3.35) | 1.48 (0.71-3.10) | 1.10 (0.41-2.95) | 0.95 (0.35-2.53) |
| 13-24 | 2.16 (1.21-3.85) | 2.10 (1.13-3.91) | 0.91 (0.35-2.38) | 0.83 (0.31-2.20) |
| 25-36 | 1.30 (0.66-2.59) | 1.21 (0.58-2.54) | 1.31 (0.59-2.92) | 1.23 (0.55-2.76) |
| ≥ 37 | Referent | Referent | Referent | Referent |
| P6M sexual agreement with partner |  |  |  |  |
| Monogamous agreement | Referent | Referent | Referent | Referent |
| Non-monogamous agreement | 1.14 (0.63-2.06) | 1.19 (0.63-2.27) | 1.72 (0.77-3.83) | 1.78 (0.80-3.94) |
| No agreement | 2.07 (1.11-3.83) | 2.22 (1.13-4.36) | 2.36 (0.91-6.15) | 2.26 (0.88-5.84) |
| Childhood sexual abuse | 1.76 (1.08-2.88) | 1.84 (1.10-3.09) | 1.39 (0.74-2.58) | 1.46 (0.76-2.77) |
| P6M transactional sex |  |  |  |  |
| Providing | 2.12 (0.93-4.84) | 1.87 (0.82-4.25) | 4.19 (1.81-9.68) | 3.48 (1.65-7.35) |
| Soliciting | 1.84 (0.66-5.12) | 1.77 (0.67-4.64) | 4.50 (1.70-11.90) | 3.76 (1.68-8.44) |
| Anxious symptomatology<sup>e</sup> | 1.84 (1.16-2.90) | 1.78 (1.12-2.83) | 1.06 (0.58-1.95) | 1.10 (0.58- 2.11) |
| Depressive symptomatology<sup>e</sup> | 1.45 (0.90-2.33) | 1.34 (0.82-2.19) | 1.80 (1.06-3.06) | 1.62 (0.94-2.79) |
| P6M problematic alcohol use<sup>f</sup> | 1.19 (0.79-1.81) | 1.12 (0.73-1.69) | 1.07 (0.61-1.88) | 0.96 (0.52-1.76) |
| P6M unregulated drug use<sup>g</sup> | 1.80 (1.12-2.87) | 1.70 (1.05-2.76) | 2.29 (1.31-3.99) | 2.25 (1.25-4.07) |
| P6M unregulated opioid use | 1.26 (0.44-3.59) | 1.13 (0.40-3.15) | 2.04 (0.58-7.14) | 1.80 (0.59-5.54) |
| P6M injection drug use | 5.50 (2.95-10.27) | 5.68 (2.92-11.02) | 4.88 (2.00-11.91) | 5.04 (2.01-12.66) |

<sup>a</sup> Missingness covariate indicator method is used, but “Missing” results are not presented for simplicity.
<sup>b</sup> Stabilized IPCW are used to account for loss to follow-up.
<sup>c</sup> All multivariable models adjust for age, city, sexual orientation, gender, ethnocultural group, and educational attainment.
<sup>d</sup> Trans man was included in “Other” due to low counts.
<sup>e</sup> Refers to 1-week period prior to interview.
<sup>f</sup> Refers to a grouped AUDIT-C score > 4.
<sup>g</sup> Excluding opioids.
IPV=intimate partner violence; PR=prevalence ratio; aPR=adjusted prevalence ratio; IPCW=inverse probability censoring weights; P6M=past six months.

### Determinants of intimate partner violence perpetration in the past 6 months

Baseline reporting of prior experience (aPR 2.78, 95% CI: 1.54-5.02) and perpetration (aPR 6.11, 95% CI: 3.46-10.79) of IPV were associated with an increased risk of P6M perpetration. We found a positive association between P6M IPV perpetration and the following covariates: Indigenous identity, lower education level, no sexual agreement with their partner, and unregulated substance use (injection and non-injection). Bisexual identity, depressive symptomatology, and history of selling sex were also associated with an increased risk of P6M IPV perpetration (Table 3).

### Temporal trends of intimate partner violence and COVID-19 restrictions

After adjusting for covariates, the impact of periods of COVID-19 physical-distancing on P6M IPV experience (aPR 0.90, 95% CI: 0.60-1.34) and perpetration (aPR 0.82, 95% CI: 0.48-1.38) were inconclusive and had large uncertainties (Table 4). Overall, we found P6M IPV to be generally stable across all three cities over the study period (Figure 1). This was also the case in Toronto after removing an outlier observation (Supplementary Figure S2).

**Figure 1.**
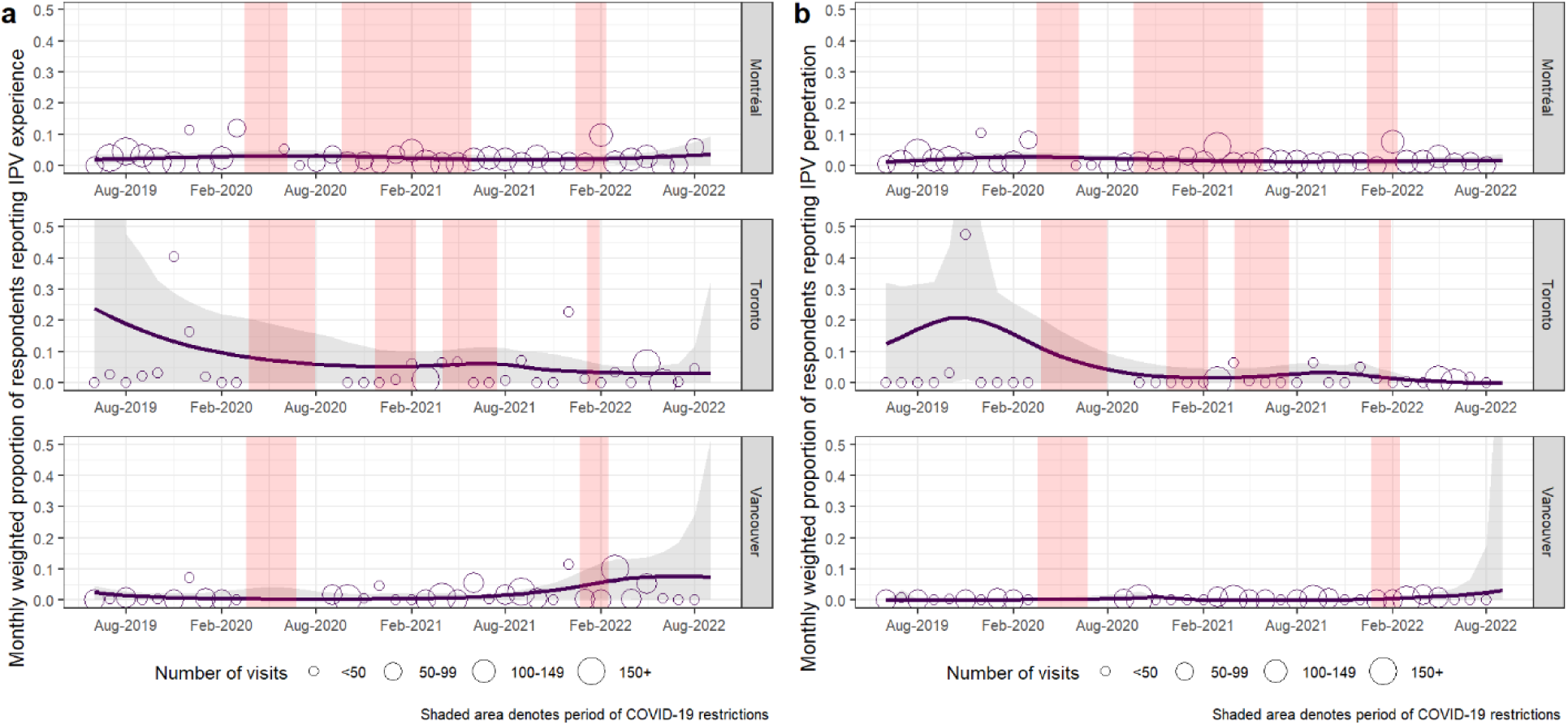
Monthly proportion of physical and/or sexual intimate partner violence (a) experience or (b) perpetration among partnered *Engage* participants in Montréal, Toronto, and Vancouver (respondent-driven sampling and inverse probability of censoring-adjusted, August 2019–August 2022). Lines represent the overall temporal trends and the grey ribbon around is the 95% confidence interval. The points represent the monthly fraction of respondents reporting IPV with the size being relative to the number of participants that month. Finally, periods of physical-distancing restrictions correspond to the vertical red bars. Knots were located at the following dates for P6M IPV experience in Montréal (Aug 5, 2019, Oct 29, 2020, May 18, 2021), Toronto (Apr 15, 2021, Jun 30, 2021, Sep 20, 2021), and Vancouver (Mar 2, 2020, Apr 14, 2021, Apr 12, 2022). Knots were located at the following dates for P6M IPV perpetration Montréal (Aug 5, 2019, May 18, 2021, Jun 12, 2022), Toronto (Sep 10, 2019, Apr 15, 2021, Sep 20, 2021), and Vancouver (Sep 28, 2020, Oct 30, 2020, Apr 4, 2021, Sep 7, 2021). Four knot locations were determined for P6M IPV perpetration in Vancouver due to uninterpretable results in the tails.

**Table 4.** Association between periods of COVID-19 pandemic health restrictions and incident experience or perpetration of intimate partner violence among partnered *Engage* participants in Montréal, Toronto, and Vancouver (crude and adjusted, 2017–2022).

|  | Period of COVID-19 physical distancing restrictions |  |
| --- | --- | --- |
|  | PR (95% CI)<sup>a</sup> | aPR (95% CI)<sup>a, b</sup> |
| IPV experience<sup>c</sup> |  |  |
| Physical and/or sexual | 0.94 (0.65-1.37) | 0.90 (0.60-1.34) |
| Physical | 1.04 (0.70-1.53) | 0.99 (0.63-1.54) |
| Sexual | 1.04 (0.46-2.37) | 0.97 (0.46-2.06) |
| IPV perpetration<sup>c</sup> |  |  |
| Physical and/or sexual | 0.93 (0.58-1.52) | 0.82 (0.48-1.38) |
| Physical | 0.87 (0.53-1.43) | 0.74 (0.43-1.26) |
| Sexual | 1.89 (0.55-6.52) | 1.86 (0.49-7.04) |

<sup>a</sup> Stabilized IPCW are used to account for loss to follow-up.
<sup>b</sup> All multivariable models adjust for age, city, sexual orientation, gender, ethnocultural group, and educational attainment.
<sup>c</sup> Refers to the 6-month period prior to interview.
IPCW=inverse probability censoring weight; IPV=intimate partner violence; PR=prevalence ratio; aPR=adjusted prevalence ratio.

### Sensitivity analysis

Including verbal IPV in our outcome definition did not qualitatively change most associations. However, when including all three forms of IPV in our definition (i.e., physical, sexual, or verbal), the association between education and income with P6M IPV experience and perpetration of IPV were closer to the null (Supplementary Table S5). Moreover, relationship duration was less robust of a predictor of P6M IPV experience and transactional sex was less important for P6M IPV perpetration, when including verbal IPV in the outcome. When we evaluated the impact of COVID-19 physical distancing on any physical, sexual, or verbal IPV, we observed a lower likelihood of verbal IPV experience and perpetration during COVID-19 (Supplementary Table S6).

## Discussion

Close to 1 in 3 partnered GBM from Canada’s three largest cities reported ever experiencing physical and/or sexual IPV in their lifetime, and 16% reported ever perpetrating it. Over five years of follow-up visits (2017-2022), 7% of GBM in our sample reported experiencing IPV in the P6M at least once, and 4% reported perpetrating it. During follow-up, younger and multiply marginalized (e.g., Indigenous and/or low educational attainment) individuals were at increased risk of P6M IPV experience and perpetration.

In all three cities the prevalence of P6M experience and perpetration of IPV was stable across time. The associations between periods of COVID-19 physical distancing and IPV experience and perpetration were inconclusive as the estimates had large uncertainties. Together, these results call for attention to the high burden of IPV among GBM, particularly given that estimates are on par with that of male-perpetrated IPV against women.^46,47^ For example, the 2018 WHO report on IPV against women found that 25% (uncertainty interval [UI] 14-41%) of ever partnered women aged 15 years or older in North America have experienced physical and/or sexual IPV in their lifetime while 6% (UI: 4-9%) experienced it in the past year.^1^

Our study identified several determinants associated with IPV experience and perpetration over time. These include reporting IPV experience or perpetration at baseline, lower education level, and substance use, consistent with preceding studies among GBM.^4,6,9^ In addition to their sexual orientation and/or gender identity, GBM who are living with HIV, identify as Indigenous, and/or experienced CSA face increased risks of experiencing IPV.

This lends support to the syndemic theory^48^ as marginalized identities and health states co-occur and increase the risk of IPV experience and perpetration. Focused interventions and support services should account for these intersecting and complex factors that contribute to IPV among GBM. As suggested by others^49^, we recommend that supportive programs include training for IPV among GBM using an intersectional perspective to address the vast challenges that GBM may face.

Of note, our analysis lacked precision to draw a strong association between IPV experience and problematic alcohol use, as observed in previous cross-sectional studies.^15,16,50^ This could be attributed to sample differences; previous studies have relied on small samples of ethnically diverse GBM^15,50^ or used a different questionnaire to approximate problematic alcohol use.^16^ In contrast, our sample was predominantly white with a lower prevalence of problematic drinking.

Cross-sectional studies using convenience samples of GBM have reported an association between IPV and COVID-19 pandemic restrictions.^51,52^ Our main analyses, however, did not detect an important association between IPV and COVID-19 restrictions in our sample. This discrepancy underscores the complexity of estimating rapid changes in IPV using a 6-month recall period. In addition, it is possible that pandemic restrictions had an impact on the frequency or severity of IPV, but this information was not collected in Engage. Finally, it is possible that a larger fraction of GBM did not live with their main partner and were thus affected differently by restrictions.

Several limitations should be addressed. First, self-reported IPV measures are subject to underreporting due to the stigma associated with IPV and social desirability bias.^22,25^ This is especially pertinent when interpreting results on perpetration. Second, the items used in the main questionnaires did not capture the frequency or severity of IPV. Additionally, we lacked gender information of the perpetrator or victim of IPV when not experienced or perpetrated by the participant. This hinders our understanding of IPV among bisexual men who may have been in relationships with people of a different gender. Third, our definition of IPV focused only on physical and sexual violence and did not consider other forms of violence (e.g., coercive control, technological, or financial). However, including verbal IPV in a sensitivity analysis did not majorly alter our conclusions. Fourth, GBM living outside of large metropolitan regions can experience different forms and rates of IPV^27^ and our data may not be able to speak to these experiences given our focus on GBM living in urban centres. Fifth, as is the case with all RDS surveys of hard-to-reach populations,^34^ we assume that all participants were able to accurately report their network size. Finally, our analysis of determinants of IPV could have been affected by residual confounding and/or reverse causality for some variables.

Strengths of our study include a large sample, collected using RDS in Canada’s three largest cities, with detailed longitudinal information to estimate prevalence and incidence of recent IPV over time.

## Conclusion

Consistent with prior studies, we found a high prevalence of both IPV experience and perpetration among GBM living in the three largest urban centers of Canada. We found that multiple social determinants of health and health states (e.g., lower income, living with HIV, prior CSA) and marginalized identities (e.g., race/ethnicity, unregulated substance use) could increase the risk of experiencing IPV. The high prevalence of IPV experience and perpetration among GBM underscores the need for targeted research and interventions addressing co-occurring health states, identities, and syndemic factors associated with increased IPV risk among GBM. Ultimately, these findings emphasize the importance of prevention and screening of at-risk individuals, linkage to and scale-up of GBM-dedicated violence prevention resources, and addressing the pervasive discrimination faced by this population.

## Contributions

S.J., M.M-G., and J.C. conceived of and designed the study. Formal data analysis was performed by S.J., with support from M.M-G., J.L.F.A., and M.D. S.J. wrote the initial draft. Overall supervision for this project was provided by M.M-G. and J.C. All authors read, approved, and jointly shared responsibility to submit the final article for publication. The Engage Cohort Study is led by the following principal investigators in Toronto by D.G. and T.A.H, in Montreal by J.C. and G.L.; and in Vancouver by J.J., N.J.L., and D.M.M.

## Data Availability

All data produced in the present study are available upon reasonable request to the authors.

## Acknowledgments

Data from this work comes from the Engage study. The principal investigators of the Engage study are Joseph Cox and Gilles Lambert (Montréal), Jody Jollimore, Nathan J. Lachowsky and David M. Moore (Vancouver), and Daniel Grace and Trevor A. Hart (Toronto). The authors thank the Engage/Momentum II study participants, office staff, and community engagement committee members, as well as their community partner agencies. The authors also wish to acknowledge the support of any Co-Is, Co-PIs, or other collaborators who were part of the process of preparing components of this manuscript/work, but who were not eligible for or declined authorship and their contribution(s) to the work presented here. More information about the Engage Cohort Study can be found here: https://www.engage-men.ca/.

