## Supplemental Files for "Prevalence, determinants, and trends in the experience and perpetration of intimate partner violence among a cohort of gay, bisexual, and other men who have sex with men in Montréal, Toronto, and Vancouver, Canada (2017-2022)"

### Supplementary Materials

### Appendix I: Supplementary methods

**Table S1.** Selected measures and details

| Domain | Measure |
|---|---|
| Socio-demographic characteristic | Participants reported their age, sexual orientation, gender identity, ethnocultural group, immigration status, education level and annual income. These characteristics were categorized as per prior Engage studies.[1–4] |
| HIV status | HIV serostatus was ascertained using fourth generation testing (detection of HIV antibodies and p24 antigen) and a confirmation test (e.g., Western blot analysis). |
| Partnership characteristics | We categorized participants' partnership status (married/common-law, dating/lover, divorced/separated/widowed, single), relationship duration (≤6, 7–12, 13–24, 25–36, and ≥37 months), sexual agreement type (monogamous, non-monogamous, no agreement), and whether they were cohabitating. |
| Childhood sexual abuse (CSA) | CSA was measured using the five-item sexual abuse sub-scale of the Childhood Trauma Questionnaire-Short Form (CTQ-SF) at the baseline visit.[5] The response scale ranged from "never" (1) to "very often" (5). Participants who responded "rarely", "sometimes", "often", or "very often" to any question were coded as having experienced CSA. |
| Transactional sex | Participants reported whether they received money, drugs, or goods in exchange for sex in the past six months (P6M). Participants were also asked whether they had solicited transactional sex by providing money, drugs, or goods in exchange for sex in the P6M. |
| Mental health disorders | Anxiety and depression were assessed via the 14-item Hospital Anxiety and Depression Scale (HADS) which assessed symptomatology in the past week.[6] Individual items were scored between 0-3, and sub-scales for anxiety and depression were summed respectively to obtain a total score. As per previous cutoff scores, a score of ≥8 was coded as screening positive for depression while a score of ≥11 was coded as screening positive for anxiety (dichotomized). |
| Problematic alcohol use | Alcohol use was measured using the three-item Alcohol Use Disorders Identification Test Consumption (AUDIT-C).[7] The questions asked participants the frequency of their alcohol use, how many drinks were consumed on a typical day, and the frequency of consuming ≥6 drinks on one occasion. Each question was scored between 0-4 where a total score >4 indicates problematic drinking. |
| Unregulated substance use | We constructed three binary variables for unregulated substances in the P6M: use of non-opioid recreational drugs (cocaine, amphetamines, inhalants, sedatives, and hallucinogens), use of opioids, and any intravenous drug use. Cannabis use was not included in our definition of unregulated substances as cannabis use is legal in Canada. We did not classify opioids as recreational drugs as people who use opioids have distinct behaviours from those who engage in other illicit drugs (e.g., chemsex).[8] This distinction is consistent with prior studies among GBM in Canada and elsewhere.[8–10] |
| COVID-19 pandemic restrictions | To evaluate whether IPV changed during COVID-19 pandemic restrictions, an indicator variable was created to differentiate study visits that occurred during periods of COVID-19 physical-distancing restrictions. For each city, the start of the pandemic restrictions period was defined as the date of mandated work-from-home orders, prohibited indoor gatherings, or imposed curfews,[11] whichever was earlier. This period ended when indoor gatherings of six or more people were permitted or when curfews were lifted, whichever was later (plus six months to account for the six-month recall period). |

**Table S2.** STROBE-RDS Checklist.

In the table below, page numbers refer to the main text of the manuscript. Tables and figures without prefixes refer to those of the manuscript. The following prefixes are used for other sources: “P” refers to page numbers from prior publications, superscript numbers refer to the reference, and “S” refers to page numbers of supplementary files.

| | Item No | Recommendation | Page No |
|---|---|---|---|
| **Title and abstract** | 1 | (*a*) Indicate “respondent-driven sampling" in the title or abstract | 1 |
| | | (*b*) Provide in the abstract an informative and balanced summary of what was done and what was found | 2 |
| **Introduction** | | | |
| Background/rationale | 2 | Explain the scientific background and rationale for the investigation being reported | 3 |
| Objectives | 3 | State specific objectives, including any prespecified hypotheses | 3 |
| **Methods** | | | |
| Study design | 4 | (*a*) Present key elements of study design early in the paper | 4 |
| | | (*b*) State why RDS was chosen as the sampling method | 4 |
| Setting | 5 | (*a*) Describe the setting, locations, and relevant dates, including periods of recruitment, and data collection | 4, S4 |
| | | (*b*) Describe formative research findings used to inform RDS study | P[1]S1 |
| Participants | 6 | (a) Give the eligibility criteria, and the sources and methods of selection of participants. Describe how participants were trained/ instructed to recruit others, number of coupons issued per person, any time limits for referral | P[1]S3, P[12]S1 |
| | | (b) Describe methods of seed selection and state number at start of study and number added later | P[1]S1, P[12]S1 |
| | | (c) State if there was any variation in study procedures during data collection (e.g., changing numbers of coupons per recruiter, interruptions in sampling, or stopping recruitment chains) | n/a |
| | | (d) Report wording of personal network size question(s) | P[1]S3 |
| | | (e) Describe incentives for participation and recruitment | 4, P[1]S2 |
| Variables | 7 | (a) If applicable, clearly define all outcomes, correlates, predictors, potential confounders, effect modifiers, and diagnostic criteria | 4-5 |
| | | (b) State how recruiter-recruit relationship was tracked | P[1]S4 |
| Data sources/ measurement | 8 | *(a)* For each variable of interest, give sources of data and details of methods of measurement. Describe comparability of measurement methods if there is more than one group | 4-5 |
| | | *(b)* Describe methods to assess eligibility and reduce repeat enrollment (e.g. coupon manager software, biometrics) | P[1]S2-3 |
| Bias | 9 | Describe any efforts to address potential sources of bias | 6, S4 |
| Study size | 10 | Explain how the study size was arrived at | P[2]530 |
| Quantitative variables | 11 | Explain how quantitative variables were handled in the analyses. If applicable, describe which groupings were chosen and why | 4,6 |
| Statistical methods | 12 | (a) Describe all statistical methods, including those to account for sampling strategy (e.g. the estimator used) and, if applicable, those used to control for confounding | 6 |
| | | (b) State data analysis software, version number and specific analysis settings used | 6, S4 |
| | | (c) Describe any methods used to examine subgroups and interactions | 6 |
| | | (d) Explain how missing data were addressed | 6, Table 2 |

| | | | |
|---|---|---|---|
| | | (e) Describe any sensitivity analyses | 6 |
| | | (f) Report any criteria used to support statements on whether estimator conditions or assumptions were appropriate | n/a |
| | | (g) Explain how seeds were handled in analysis | n/a |
| **Results** | | | |
| Participants | 13 | (a) Report the numbers of individuals at each stage of the study —e.g., numbers potentially eligible, examined for eligibility, confirmed eligible, included in the study, and analyzed | Fig S1, P[12]S1 |
| | | (b) Give reasons for non-participation at each stage (e.g., not eligible, does not consent, decline to recruit others) | n/a |
| | | (c) Consider use of a flow diagram | Fig S1 |
| | | (d) Report number of coupons issued and returned | P[1]S3, P[12]S1 |
| | | (e) Report number of recruits by seed and number of RDS recruitment waves for each seed. Consider showing graph of entire recruitment network | P[2] Fig S2-S4 |
| | | (f) Report recruitment challenges (e.g. commercial exchange of coupons, imposters, duplicate recruits) and how addressed | n/a |
| | | (g) Consider reporting estimated design effect for outcomes of interest | n/a |
| Descriptive data | 14 | (a) Give characteristics of study participants (e.g., demographic, clinical, social) and, if applicable, information on correlates and potential confounders. Report unweighted sample size and percentages, estimated population proportions or means with estimated precision (e.g., 95% confidence interval) | 6-7, Table 1 |
| | | (b) Indicate number of participants with missing data for each variable of interest | n/a |
| | | (c) Summarise follow-up time (eg, average and total amount) | P[12]S1 |
| Outcome data | 15 | If applicable, report number of outcome events or summary measures | 7 |
| Main results | 16 | (a) Give unadjusted and study design adjusted estimates and, if applicable, confounder adjusted estimates and their precision (e.g., 95% confidence intervals). Make clear which confounders were adjusted for and why they were included | Table 2, Table 3 |
| | | (b) Report category boundaries when continuous variables were categorised | Table 2, Table 3 |
| | | (c) If adjustment of primary outcome leads to marked changes, report information on factors influencing the adjustments (e.g. personal network sizes, recruitment patterns by group, key confounders) | n/a |
| Other analyses | 17 | Report other analyses done—e.g., analyses of subgroups and interactions, sensitivity analyses, different RDS estimators and definitions of personal network size | Table S1, Table S2, Table S3 |
| **Discussion** | | | |
| Key results | 18 | Summarise key results with reference to study objectives | 8 |
| Limitations | 19 | Discuss limitations of the study, taking into account sources of potential bias or imprecision. Discuss both direction and magnitude of any potential bias | 9 |
| Interpretation | 20 | Give a cautious overall interpretation of results considering objectives, limitations, multiplicity of analyses, results from similar studies, and other relevant evidence | 8-9 |
| Generalisability | 21 | Discuss the generalisability (external validity) of the study results | 9 |
| **Other information** | | | |
| Funding | 22 | Give the source of funding and the role of the funders for the present study and, if applicable, for the original study on which the present article is based | 11 |

**a) Survey questions for the primary outcome of physical and/or sexual intimate partner violence (IPV) experience and perpetration, and for the outcome of verbal IPV for sensitivity analysis**

The questionnaire used in the *Engage Cohort Study* asked participants the following questions on their experience and perpetration of physical and/or verbal intimate partner violence (IPV):

(1) *Have you been hit, kicked, or slapped by a lover or boyfriend in the past 6 months?*
(2) *Have you been sexually abused or raped by a lover or boyfriend in the past 6 months?*
(3) *Have you hit, kicked, or slapped a lover or boyfriend in the past 6 months?*
(4) *Have you sexually abused or raped a lover or boyfriend in the past 6 months?*

These questions were adapted from prior studies using a modified version of the Conflict Tactics Scale[13,14]. Participants who affirmatively answered either (1) or (2) were considered to have experienced P6M IPV, and those who affirmatively answered either (3) or (4) were considered to have perpetrated P6M IPV.

For the experience or perpetration of verbal IPV, the questions used were:

(1) *Have you been insulted or verbally abused by a lover of boyfriend in the past 6 months?*
(2) *Have you insulted or verbally abused a lover or boyfriend in the past 6 months?*

Participants who positively responded (1) were considered to have experienced P6M verbal IPV, and those who positively responded (2) were considered to have perpetrated verbal IPV in the P6M.

**b) Computation of inverse probability of censoring weights**

We calculated inverse probability of censoring weights (IPCW) to reduce bias resulting from loss to follow-up (LTFU) of participants, following the method described by Willems et al[15]. Covariate imbalance between participants who had follow-up data and who had missed a given visit was assessed using standardized mean difference (SMD) for each follow-up visit in each city. Covariates were included in the LTFU model if SMD > 0.1. The covariates considered and assessed for imbalance include all measures described in the main text.

**c) List of R packages used**

***Data cleaning***

Handcock MS, Gile KJ, Fellows IE, Neely WW. **RDS**: Respondent-Driven Sampling. 2023 [cited 2023 May 24]. Available from: https://cran.r-project.org/web/packages/RDS/index.html

Spinu V, Grolemund G, Wickham H, Vaughan D, Lyttle I, Costigan I, et al. **lubridate**: Make Dealing with Dates a Little Easier. 2022 [cited 2023 May 24]. Available from: https://cran.r-project.org/web/packages/lubridate/index.html1.

Wickham H, François R, Henry L, Müller K, Vaughan D, Software P, et al. **dplyr**: A Grammar of Data Manipulation. 2022 [cited 2023 May 24]. Available from: https://cran.r-project.org/web/packages/dplyr/index.html

Wickham H, Miller E, Smith D. **haven**: Import and Export "SPSS", "Stata" and "SAS" Files. 2022 [cited 2023 May 24]. Available from: https://cran.r-project.org/web/packages/haven/index.html

Zeileis A, Grothendieck G, Ryan JA, Ulrich JM, Andrews F. **zoo**: S3 Infrastructure for Regular and Irregular Time Series (Z's Ordered Observations). 2023 [cited 2023 May 24]. Available from: https://cran.r-project.org/web/packages/zoo/index.html

***Data analysis***

Barnier J, Briatte F, Larmarange J. **questionr**: Functions to Make Surveys Processing Easier. 2022 [cited 2023 May 24]. Available from: https://cran.r-project.org/web/packages/questionr/index.html

Højsgaard S, Halekoh U, Yan J, Ekstrøm CT. **geepack**: Generalized Estimating Equation Package. 2022 [cited 2023 May 24]. Available from: https://cran.r-project.org/web/packages/geepack/index.html

Lumley T. **survey**: Analysis of Complex Survey Samples. 2021 [cited 2023 May 24]. Available from: https://cran.r-project.org/web/packages/survey/index.html

***Data visualization***

Bates DM, Venables WN. **splines**: Regression Spline Functions and Classes. 2021 [cited 2023 May 24]. Available from: https://stat.ethz.ch/R-manual/R-devel/library/splines/html/splines-package.html

Garnier S, Ross N, Rudis B, Sciaini M, Camargo AP, Scherer C. **viridis**: Colorblind-Friendly Color Maps for R. 2023 [cited 2023 May 24]. Available from: https://cran.r-project.org/web/packages/viridis/index.html

Kassambara A. **ggpubr**: "ggplot2" Based Publication Ready Plots. 2020 [cited 2023 May 24]. Available from: https://cran.r-project.org/web/packages/ggpubr/index.html

Murrell P. **grid**: The Grid Graphics Package. 2021 [cited 2023 May 24]. Available from: https://stat.ethz.ch/R-manual/R-devel/library/grid/html/00Index.html

Rich B. **table1**: Tables of Descriptive Statistics in HTML [Internet]. 2021 [cited 2023 May 24]. Available from: https://cran.r-project.org/web/packages/table1/index.html

Wickham H, Chang W, Henry L, Pedersen TL, Takahashi K, Wilke C, et al. **ggplot2**: Create Elegant Data Visualisations Using the Grammar of Graphics. 2022 [cited 2023 May 24]. Available from: https://cran.r-project.org/web/packages/ggplot2/index.html

**Figure S1:** Participant flowchart

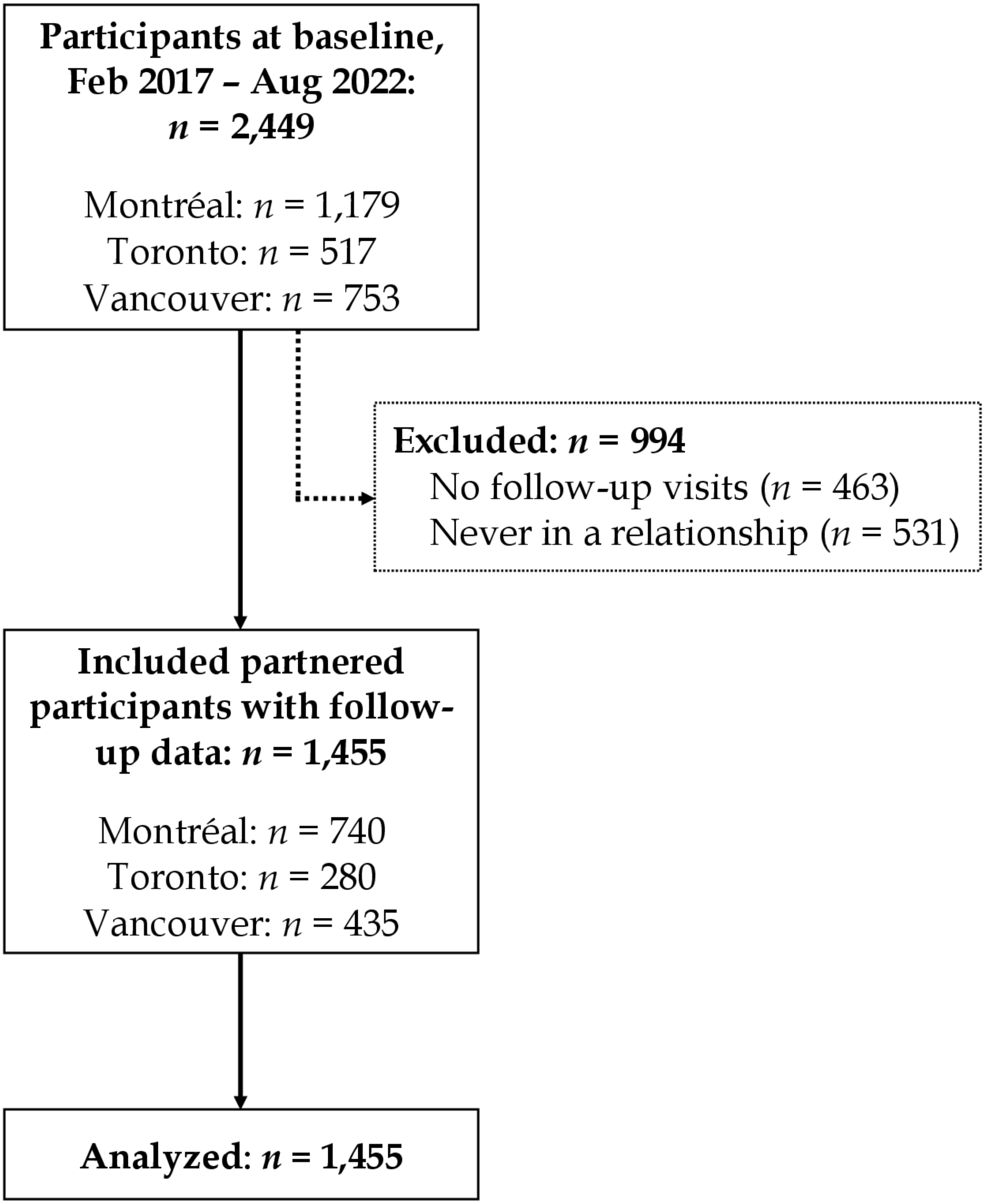

### Appendix II: Supplementary results, prevalence of verbal intimate partner violence experience and perpetration among partnered gay, bisexual, and other men who have sex with men (GBM)

**Table S3.** Experience and perpetration of verbal, physical, and/or sexual intimate partner violence among partnered *Engage* participants with follow-up in Montréal, Toronto, and Vancouver (crude and respondent-driven sampling-adjusted, 2017-2019).

| | **Overall** (n=1,455) | **Montréal** (n=740) | | **Toronto** (n=280) | | **Vancouver** (n=435) | |
|---|---|---|---|---|---|---|---|
| | **Crude %** | **Crude %** | **RDS % (95%CI)** | **Crude %** | **RDS % (95%CI)** | **Crude %** | **RDS % (95%CI)** |
| **IPV variable**[a] | | | | | | | |
| Lifetime experience of any IPV[b] | 50.8 | 48.8 | 45.3 (38.8-51.9) | 53.9 | 35.6 (26.4-44.7) | 52.2 | 38.6 (30.6-46.6) |
| Missing | 0.4 | 0.7 | 2.1 (0.1-4.1) | 0.4 | 0.1 (0.0-0.5) | 0.0 | 0.0 (0.0-0.0) |
| Any verbal IPV | 46.3 | 43.6 | 39.4 (33.0-45.9) | 50.7 | 31.0 (22.0-40.0) | 47.8 | 36.5 (28.6-44.4) |
| Missing | 0.8 | 1.4 | 3.9 (0.8-7.0) | 0.4 | 0.1 (0.0-0.5) | 0.2 | 0.2 (0.0-0.4) |
| Lifetime perpetration of any IPV[b] | 36.2 | 35.8 | 31.2 (25.4-37.0) | 33.9 | 22.1 (14.4-29.7) | 38.4 | 27.1 (19.8-34.3) |
| Missing | 0.7 | 0.8 | 0.2 (0.1-4.2) | 0.4 | 0.1 (0.0-0.5) | 0.7 | 0.9 (0.0-3.7) |
| Any verbal IPV | 32.8 | 32.3 | 27.7 (22.3-33.2) | 31.1 | 20.9 (13.3-28.4) | 34.7 | 23.6 (16.6-30.6) |
| Missing | 0.9 | 1.2 | 4.0 (6.7-7.3) | 0.4 | 0.1 (0.0-0.5) | 0.7 | 0.9 (0.0-3.7) |

[a] Refers to lifetime exposure to covariate.

[b] Includes any verbal, physical, and/or sexual IPV.

IPV=intimate partner violence; RDS=respondent-driven sampling; 95%CI=95% confidence interval.

**Table S4.** Incidence of self-reported verbal, physical, and/or sexual intimate partner violence among partnered *Engage* participants with follow-up in Montréal, Toronto, and Vancouver (crude and respondent-driven sampling-adjusted, 2017–2022).

| | | **Experience of verbal, physical, and/or sexual IPV** | | | | | | | |
|---|---|---|---|---|---|---|---|---|---|
| | | **Overall** (n=1,455)[a] | | **Montréal** (n=740)[a] | | **Toronto** (n=280)[a] | | **Vancouver** (n=435)[a] | |
| **Lifetime IPV**[b] | **Incident P6M IPV**[b] | **n (%)** | **Mean # of follow-up visits** | **n (%)** | **Adjusted %**[d] | **n (%)** | **Adjusted %**[d] | **n (%)** | **Adjusted %**[d] |
| Yes | Yes | 183 (12.6) | 4.2 | 101 (13.6) | 11.5 | 29 (10.4) | 6.4 | 49 (11.3) | 6.0 |
| | No | 454 (31.2) | 3.5 | 21.7 (29.3) | 26.8 | 104 (37.1) | 25.5 | 117 (26.9) | 22.4 |
| No | Yes | 62 (4.3) | 4.4 | 33 (4.5) | 2.9 | 16 (5.7) | 6.6 | 11 (2.5) | 5.8 |
| | No | 571 (39.2) | 3.9 | 312 (42.2) | 44.1 | 91 (32.5) | 31.8 | 156 (35.9) | 47.9 |
| | | **Perpetration of verbal, physical, and/or sexual IPV** | | | | | | | |
| | | **Overall** (n=1,455)[a] | | **Montréal** (n=740)[a] | | **Toronto** (n=280)[a] | | **Vancouver** (n=435)[a] | |
| **Lifetime IPV**[c] | **Incident P6M IPV**[c] | **n (%)** | **Mean # of follow-up visits** | **n (%)** | **Adjusted %**[d] | **n (%)** | **Adjusted %**[d] | **n (%)** | **Adjusted %**[d] |
| Yes | Yes | 121 (8.3) | 4.3 | 66 (8.9) | 6.7 | 21 (7.5) | 3.6 | 33 (7.6) | 2.9 |
| | No | 331 (22.7) | 3.5 | 165 (22.3) | 18.2 | 63 (22.5) | 16.8 | 90 (20.7) | 17.0 |
| No | Yes | 42 (2.9) | 4.1 | 27 (3.7) | 2.4 | 12 (4.3) | 5.6 | 2 (0.5) | 0.6 |
| | No | 773 (53.1) | 3.9 | 404 (54.6) | 58.0 | 144 (51.4) | 44.2 | 206 (47.4) | 61.3 |

[a] Missing, poor data quality, and "prefer not to answer" responses removed when applicable but included in the denominator.
[b] IPV experience.
[c] IPV perpetration.
[d] Proportions weighted using RDS-II weights.

IPV=intimate partner violence; GBM=gay, bisexual, and other men who have sex with men.

**Table S5.** Association between verbal, physical, and/or sexual intimate partner violence (IPV) experience or perpetration, socio-demographic, partnership characteristics, mental health, and substance use during follow-up among partnered *Engage* participants in Montréal, Toronto, and Vancouver (crude and adjusted, 2017–2022).

| | Verbal, physical, and/or sexual IPV experience in the past 6 months[a] | | Verbal, physical, and/or sexual IPV perpetration in the past 6 months[a] | |
|---|---|---|---|---|
| | **PR (95% CI)** | **aPR (95% CI)[b]** | **PR (95% CI)** | **aPR (95% CI)[b]** |
| Reported IPV experience at baseline | 3.35 (2.49-4.52) | 3.27 (2.43-4.42) | 3.44 (2.34-5.05) | 3.16 (2.14-4.66) |
| Reported IPV perpetration at baseline | 2.96 (2.28-3.85) | 2.89 (2.22-3.76) | 5.93 (4.08-8.62) | 5.56 (3.82-8.11) |
| Age | | | | |
| < 30 | 1.64 (1.13-2.38) | 1.67 (1.14-2.45) | 1.39 (0.87-2.20) | 1.54 (0.95-2.50) |
| 30-44 | 1.60 (1.17-2.19) | 1.61 (1.17-2.22) | 1.60 (1.10-2.33) | 1.81 (1.23-2.67) |
| 45+ | Referent | Referent | Referent | Referent |
| City | | | | |
| Montréal | Referent | Referent | Referent | Referent |
| Toronto | 1.06 (0.75-1.48) | 1.19 (0.85-1.67) | 1.13 (0.75-1.69) | 1.38 (0.93-2.05) |
| Vancouver | 0.89 (0.65-1.20) | 1.02 (0.75-1.38) | 0.87 (0.58-1.30) | 1.12 (0.75-1.67) |
| Sexual orientation | | | | |
| Gay | Referent | Referent | Referent | Referent |
| Bisexual | 1.22 (0.73-2.03) | 1.01 (0.60-1.72) | 1.47 (0.91-2.39) | 1.21 (0.73-2.02) |
| Other | 1.28 (0.95-1.73) | 1.02 (0.73-1.42) | 1.16 (0.76-1.78) | 0.95 (0.60-1.51) |
| Gender | | | | |
| Cisgender man | Referent | Referent | Referent | Referent |
| Other[d] | 1.62 (1.12-2.34) | 1.27 (0.82-1.97) | 1.56 (0.93-2.61) | 1.19 (0.65-2.16) |
| Ethnocultural group | | | | |
| English- or French-Canadian/European | Referent | Referent | Referent | Referent |
| Aboriginal or Indigenous | 3.10 (1.79-5.38) | 2.61 (1.46-4.67) | 1.74 (0.65-4.67) | 1.09 (0.37-3.22) |
| Asian | 0.74 (0.42-1.29) | 0.69 (0.40-1.19) | 0.22 (0.08-0.60) | 0.24 (0.09-0.68) |
| Latin American | 0.85 (0.52-1.39) | 0.89 (0.56-1.41) | 0.79 (0.44-1.42) | 0.85 (0.48-1.51) |
| African/Caribbean/Black | 0.87 (0.48-1.59) | 0.86 (0.48-1.56) | 0.98 (0.48-2.00) | 0.94 (0.46-1.92) |
| Other | 0.89 (0.54-1.48) | 0.89 (0.54-1.46) | 0.85 (0.45-1.60) | 0.91 (0.49-1.69) |
| Born outside Canada | 0.65 (0.33-1.25) | 1.03 (0.46- 2.30) | 0.87 (0.62-1.24) | 1.11 (0.72-1.70) |
| Less than high school education | 1.23 (0.88-1.73) | 1.11 (0.78-1.58) | 1.73 (1.20-2.49) | 1.60 (1.11-2.30) |
| Annual income, $CAD | | | | |
| <30,000 | 1.42 (0.99-2.02) | 1.34 (0.93-1.94) | 1.11 (0.72-1.72) | 1.04 (0.67-1.64) |
| 30,000-59,999 | 1.17 (0.83-1.63) | 1.13 (0.80-1.58) | 1.12 (0.75-1.68) | 1.08 (0.71-1.63) |
| 60,000-79,999 | 1.11 (0.76-1.62) | 1.07 (0.74-1.56) | 1.08 (0.71-1.64) | 1.08 (0.70-1.65) |
| ≥80,000 | Referent | Referent | Referent | Referent |
| Living with HIV | 1.47 (1.08-2.01) | 1.80 (1.25-2.60) | 1.34 (0.87-2.08) | 1.55 (0.96-2.50) |
| Relationship Status | | | | |
| Married or common-law | 0.88 (0.64-1.21) | 0.90 (0.66-1.23) | 1.47 (0.97-2.23) | 1.48 (0.99-2.19) |
| Dating or lover | 0.91 (0.69-1.20) | 0.89 (0.68-1.16) | 1.27 (0.86-1.87) | 1.23 (0.85-1.77) |

| | | | | |
|---|---|---|---|---|
| Single, divorced, separated, or widowed | Referent | Referent | Referent | Referent |
| Cohabitating with partner | 1.07 (0.67-2.73) | 1.03 (0.81-1.29) | 1.04 (0.61-1.77) | 1.42 (1.07-1.89) |
| Relationship duration, months | | | | |
| ≤ 6 | 0.97 (0.73-1.28) | 0.88 (0.67-1.16) | 0.73 (0.50-1.07) | 0.68 (0.47-0.97) |
| 7-12 | 0.94 (0.64-1.38) | 0.83 (0.57-1.21) | 1.00 (0.64-1.54) | 0.91 (0.59-1.42) |
| 13-24 | 0.94 (0.67-1.33) | 0.86 (0.61-1.21) | 0.80 (0.52-1.22) | 0.73 (0.48-1.11) |
| 25-36 | 0.83 (0.59-1.18) | 0.78 (0.55-1.09) | 0.97 (0.68-1.40) | 0.92 (0.65-1.32) |
| ≥ 37 | Referent | Referent | Referent | Referent |
| P6M sexual agreement with partner | | | | |
| Monogamous agreement | Referent | Referent | Referent | Referent |
| Non-monogamous agreement | 1.28 (0.95-1.72) | 1.28 (0.95-1.72) | 1.60 (1.09-2.35) | 1.59 (1.10-2.31) |
| No agreement | 1.48 (1.05-2.08) | 1.51 (1.07-2.13) | 1.70 (1.10-2.63) | 1.68 (1.09-2.58) |
| Childhood sexual abuse | 1.49 (1.15-1.94) | 1.52 (1.08-2.15) | 1.10 (0.79-1.54) | 1.12 (0.80-1.57) |
| P6M sex work | | | | |
| Sells sex | 1.92 (1.29-2.85) | 1.85 (1.26-2.71) | 2.31 (1.47-3.62) | 2.21 (1.47-3.30) |
| Purchases sex | 1.55 (0.92-2.62) | 1.59 (0.99-2.56) | 2.08 (1.22-3.57) | 2.07 (1.29-3.35) |
| Anxious symptomatology[e] | 1.56 (1.25-1.96) | 1.52 (1.21-1.90) | 1.25 (0.90-1.73) | 1.22 (0.88-2.09) |
| Depressive symptomatology[e] | 1.46 (1.14-1.89) | 1.45 (1.13-1.86) | 1.47 (1.08-1.98) | 1.44 (1.07-1.94) |
| P6M problematic alcohol use[f] | 1.35 (1.08-1.70) | 1.29 (1.03-1.62) | 1.40 (1.02-1.91) | 1.34 (0.98-1.83) |
| P6M unregulated drug use[g] | 1.92 (1.50-2.47) | 1.88 (1.46-2.42) | 1.86 (1.39- 2.51) | 1.87 (1.38-2.54) |
| P6M unregulated opioid use | 1.26 (0.70-2.26) | 1.21 (0.71-2.07) | 1.16 (0.50-2.70) | 1.13 (0.55-2.33) |
| P6M injection drug use | 2.77 (1.80-4.25) | 3.15 (2.00-4.95) | 3.55 (2.20-5.73) | 3.73 (2.26-6.17) |

[a] Missingness covariate indicator method is used, but "Missing" results are not presented for simplicity.

[b] Stabilized IPCW are used to account for loss to follow-up.

[c] All multivariable models adjust for age, city, sexual orientation, gender, ethnocultural group, and educational attainment.

[d] Trans man was included in "Other" due to low counts.

[e] Refers to 1-week period prior to interview.

[f] Refers to a grouped AUDIT-C score > 4.

[g] Excluding opioids.

IPV=intimate partner violence; PR=prevalence ratio; aPR=adjusted prevalence ratio; IPCW=inverse probability censoring weights; P6M=past six months.

**Table S6.** Association between periods of COVID-19 pandemic health restrictions and incident experience or perpetration of verbal, physical, and/or sexual intimate partner violence among partnered *Engage* participants Montréal, Toronto, and Vancouver (crude and adjusted, 2017–2022).

| | Period of COVID-19 physical distancing restrictions | |
|---|---|---|
| | **PR (95% CI)[a]** | **aPR (95% CI)[b]** |
| **IPV experience[c]** | | |
| Verbal, physical, and/or sexual | 0.89 (0.74-1.06) | 0.86 (0.71-1.05) |
| Verbal | 0.93 (0.77-1.13) | 0.90 (0.74-1.11) |
| **IPV perpetration[c]** | | |
| Verbal, physical, and/or sexual | 0.68 (0.54-0.86) | 0.64 (0.50-0.82) |
| Verbal | 0.69 (0.54-0.89) | 0.65 (0.50-0.85) |

[a] Stabilized IPCW are used to account for loss to follow-up.

[b] All multivariable models adjust for age, city, sexual orientation, gender, ethnocultural group, and educational attainment.

[c] Refers to the 6-month period prior to interview.

IPV=intimate partner violence; PR=prevalence ratio; aPR=adjusted prevalence ratio; IPCW=inverse probability censoring weight.

**Appendix III: Supplementary results, longitudinal IPV trend figure without outlier**

**Figure S2.** Monthly proportion of physical and/or sexual intimate partner violence (a) experience or (b) perpetration among partnered *Engage* participants in Montréal, Toronto, and Vancouver (respondent-driven sampling and inverse probability of censoring-adjusted, August 2019–August 2022). Lines represent the overall temporal trends and the grey ribbon around is the 95% confidence interval. The points represent the monthly fraction of respondents reporting IPV with the size being relative to the number of participants that month. Note, a highly influential individual with large weights was removed from our Toronto analysis. Finally, periods of physical-distancing restrictions correspond to the vertical red bars. Knots were located at the following dates for P6M IPV experience in Montréal (Aug 5, 2019, Oct 29, 2020, May 18, 2021), Toronto (Feb 20, 2020, Apr 16, 2021, Sep 20, 2021), and Vancouver (Mar 2, 2020, Apr 14, 2021, Apr 12, 2022). Knots were located at the following dates for P6M IPV perpetration Montréal (Aug 5, 2019, May 18, 2021, Jun 12, 2022), Toronto (Sep 10, 2019, Feb 12, 2021, Aug 12, 2021), and Vancouver (Sep 28, 2020, Oct 30, 2020, Apr 4, 2021, Sep 7, 2021). Four knot locations were determined for P6M IPV perpetration in Vancouver due to uninterpretable results in the tails.

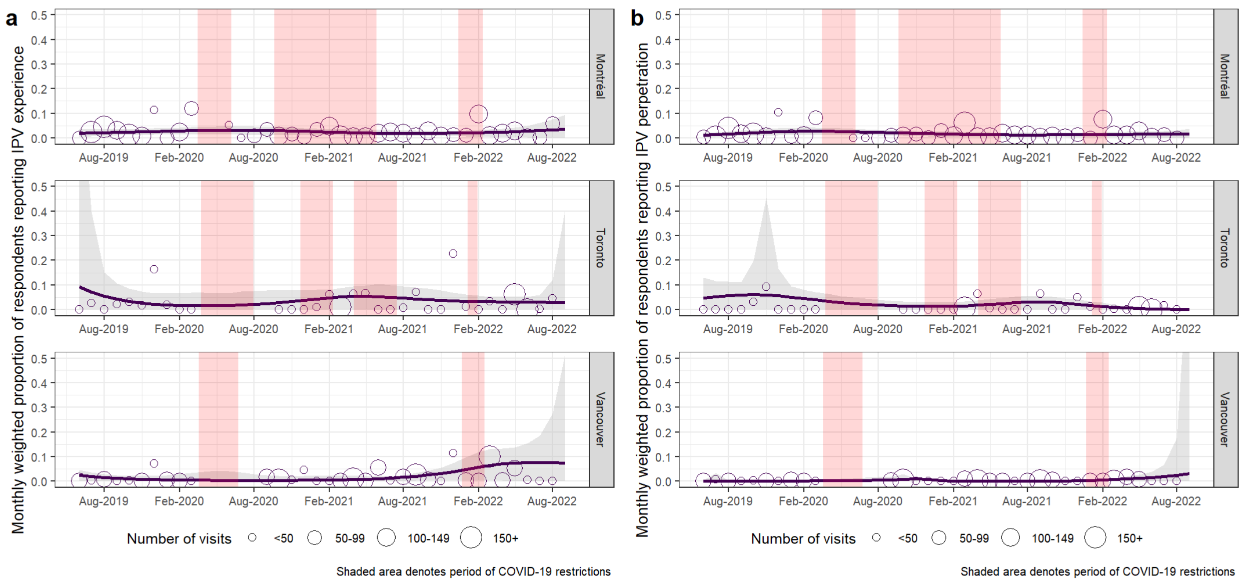
